# The Efficacy of COVID-19 Vaccines in Peritoneal Dialysis and Hemodialysis Patients

**DOI:** 10.1101/2025.04.17.25326029

**Authors:** Önder Buğra Kaynarca, Mehmet Usta, Ugur Demirpek, Alparslan Ersoy, Aslı Ceren Macunluoglu

**Affiliations:** Department of Internal Medicine, Bursa City Hospital, Bursa, Turkey; Department of Nephrology, University of Health Sciences, Bursa City Hospital, Bursa, Turkey; Department of Laboratory Medicine, Medical Microbiology Laboratory, Bursa City Hospital, Bursa, Turkey; Division of Nephrology, Department of Internal Medicine, Bursa Uludag University Faculty of Medicine, Bursa, Turkey; Department of Biostatistics, Bursa Uludag University Institute of Health Sciences, Bursa, Turkey

**Keywords:** COVID-19, chronic kidney disease, peritoneal dialysis, hemodialysis, immunisation, BioNTech, CoronaVac

## Abstract

**Introduction:** The effectiveness of COVID-19 vaccines was investigated in dialysis patients.

**Methods:** Dialysis patients and healthy individuals were included in this study. Vaccination information and post-vaccine antibody levels were obtained, and infection risk factors were examined.

**Results:** The proportion of patients receiving three doses of CoronaVac in the peritoneal dialysis group was higher than in the control and hemodialysis groups. The rate of those receiving two doses of the BioNTech vaccine in the hemodialysis group was higher than in the control group, and the rate of those receiving the combined vaccine was higher in the control group than in the peritoneal dialysis group. Post-vaccination antibody responses were higher in the control group. Administration of the combination vaccine resulted in better antibody responses. The risk of COVID-19 infection after vaccination decreased with increasing albumin levels and increased with diabetes.

**Conclusion:** Combination vaccination produced better antibody responses in maintenance dialysis patients.

This retrospective study compares the effectiveness of inactivated and mRNA COVID-19 vaccines at different applications in dialysis patients. There are very few similar studies in the literature on the dialysis population. The critical results of the study are as follows:

**Key concepts**

Response to COVID-19 vaccines in dialysis patients is lower than in the healthy population.

mRNA vaccines are more effective than inactivated vaccines.

Additional booster doses should be considered as antibody levels decrease over time.

Combining different vaccines increases protection against COVID-19.

## 1 INTRODUCTION

The novel COVID-19 infection caused by a severe acute respiratory syndrome coronavirus 2 (SARS-CoV-2) affects the elderly and individuals with severe chronic disease (chronic kidney disease [CKD], cardiovascular diseases, diabetes, chronic lung diseases, chronic liver disease and cancer, etc.). Patients with CKD usually comprise an elderly population with comorbidities such as diabetes, hypertension and cardiovascular diseases. Because of their age and weak immune systems, they are more prone to serious infectious diseases and are at higher risk of transmission than the general population [1]. In previous outbreaks, the case fatality rate in dialysis patients has always been higher than in the general population [2]. Kidney disease groups will also remain at risk until an effective treatment is found, herd immunity is achieved through vaccination, or the pandemic is over.

During the pandemic, scientists developed COVID-19 vaccines sooner than expected. After vaccines passed the necessary stages, clinicians began administering several COVID-19 vaccines approved for consecutive emergency use from December 2020. These vaccines are whole virus (inactivated or attenuated), protein subunit (protein-based), viral vector and nucleic acid (RNA and DNA) vaccines (Pfizer/BioNTech Comirnaty, 31 December 2020; SII/COVISHIELD and AstraZeneca/AZD1222 vaccines, 16 February 2021; Janssen/Ad26.COV 2.S, 12

March 2021; Moderna COVID-19 vaccine [mRNA 1273], 30 April 2021; Sinopharm COVID-19 vaccine, 7 May 2021; Sinovac-CoronaVac, 1 June 2021 and Bharat Biotech BBV152 COVAXIN vaccine, 3 November 2021) [3]. Due to uremia’s immune system suppression, post-vaccination antibody titers in dialysis patients are generally lower, and adequate antibody titers decrease over time [4]. Many studies have examined the efficacy of COVID-19 vaccines in the dialysis population. These studies include differences in vaccine type, number of vaccine doses, dose interval, administration of the same or different (combined) vaccines or measurement time of antibody response. In the present study, we aimed to investigate the efficacy of the COVID-19 vaccine in chronic dialysis patients.

## 2 METHODS

### 2.1 Study design and patients

This study retrospectively included dialysis patients and healthy individuals in a single centre and investigated persons vaccinated against SARS-Cov2 between January and August 2021. We determined from the medical records that 100 dialysis patients (50 in hemodialysis and 50 in peritoneal dialysis) and 130 healthy people were vaccinated and measured their antibody levels after vaccination. Study protocol and procedures were performed according to the 1964 Declaration of Helsinki guidelines and subsequent amendments and were approved by the local institutional review board (Date: 20 October 2021, Decision no: 2021-19/9).

We excluded individuals with a history of SARS-CoV-2 infection before vaccination and/or no antibody measurement. All patients were of Caucasian race (Turkish ethnicity). None of the patients had any history of medication or disease affecting their immune system. We divided the patients in the study population into three groups: peritoneal dialysis patients (PD group), hemodialysis patients (HD group), and a healthy control group (Control group) (Figure 1). The characteristics of the patients, including other comorbidities, were obtained from the medical records.

### 2.2 Vaccination

Vaccination centres carried out the vaccination program according to the recommendations of the Ministry of Health Science Board. The Ministry of Health announced the first COVID-19 case detected in our country on March 11, 2020. After the emergency use approval in our country, on January 14, 2021, the inactivated vaccine (CoronaVac, Sinovac Life Sciences Ltd., Beijing, China) was administered to healthcare workers and applied to the second dose of vaccines 28 days (4 weeks) later. Afterwards, the vaccination program continued gradually in high-risk groups (the elderly and those with chronic diseases). As of April 2, 2021, the vaccination program included the mRNA vaccine (BNT162b2, Pfizer-BioNTech Comirnaty vaccine). In our country, on July 1, 2021, the Ministry of Health recommended injecting the third booster vaccine for people over 50 and healthcare workers who have passed at least three months after the second vaccine. Then, the vaccination was gradually applied to all age groups. The individuals were even allowed to choose the type of vaccine (BioNTech or CoronaVac).

After the proper position of the patient was ensured, vaccination was employed on the deltoid region of the upper arm intramuscularly. For the Biontec/Pfizer vaccination, a 20 to 25-gauge needle was used. The dosage was 30 mcg (0.3 mL gray-capped vial). Cinovac Setecoject 2 mL/luer was given (employed) as 0.5 mL. All vaccines were kept in the cold chain system. No other vaccines were used at the same time.

### 2.3 Antibody measurement

Patient serum samples were collected and analysed according to the manufacturer’s standard laboratory procedures and recommendations. The antibodies to the SARS-CoV-2 spike protein were quantitatively determined on the Roche Cobas e801 analytical unit using the Roche Elecsys Anti-SARS-CoV-2 S kit (Roche Diagnostics Basel, Switzerland). Elecsys® Anti-SARS-CoV-2 S was an immunoassay for the in vitro quantitative determination of antibodies (including IgG) resulting from an adaptive humoral immune response to the SARS-CoV-2 spike (S) protein receptor-binding domain (RBD) in human serum and plasma. The assay used a recombinant protein representing the RBD of the S antigen in a double-antigen sandwich electrochemiluminescence (ECLIA) immunoassay format, favouring the detection of high-affinity antibodies against SARS-CoV-2. A value of ≥0.8 U/mL (linear range: 0.4 to 250 U/mL) was considered positive in the Elecsys® Anti-SARS-CoV-2 S assay with a clinical sensitivity of 98.8% (95% confidence interval [CI]: 98.1-99.3%) and an overall specificity of 99.98% (95% CI: 99.91-100%) in the sample cohorts. SARS-Cov2 antibody levels were measured between June and November 2021. We categorised the vaccine response according to antibody titer as follows: complete response (antibody levels >250 U/mL), suboptimal response (15 to 250 U/mL), and insufficient response (0.8 to 15 U/mL). We considered no seroconversion if the levels were <0.8 U/mL [5, 6].

### 2.4 Data analysis

Statistical analysis was performed using the SPSS (IBM Corp. Released 2012. IBM SPSS Statistics for Windows, Version 21.0, Armonk, NY: IBM Corp.). The Shapiro-Wilk test was used to assess whether the variables followed a normal distribution. Continuous variables were presented as median (minimum: maximum) or mean±standard deviation values. Categorical variables were reported as n (%). According to the normality test results, the Mann-Whitney U test or Independent samples t-test was used to compare the two groups. The Kruskal-Wallis test was used if the number of groups was more than two. Multiple comparison procedures were performed using the Dunn-Bonferroni approach to identify different group or groups after the Kruskal-Wallis test. Pearson Chi-square test, Fisher’s exact chi-square test or Fisher Freeman-Halton test were used for comparing categorical variables. Multiple logistic regression analyses via the Backwards LR procedure investigated the best independent predictor(s), which mainly affected the development of COVID-19 infection in dialysis patients. Any variable whose univariable test had a p-value less than 0.25 was accepted as a candidate for the multivariable model. Odd’s ratios (OR), 95% confidence intervals (CI) and Wald statistics for each independent variable were calculated. A p-value <0.05 was considered statistically significant.

## 3 RESULTS

In the control group, 23 subjects, 1 PD patient and 5 HD patients, were excluded from the analysis because they had COVID-19 infection, a median of 4.94 months (min-max: 0.99-8.97) before vaccination. All PD and HD patients had a complete response after vaccination. In the control group, 17 (73.9%) had a complete response, 5 (21.7%) had a suboptimal response, and 1 (4.3%) had an insufficient response.

There was a statistically significant difference between the gender and age distributions of the participants (p<0.001) (Table 1). In the subgroup analyses performed between the groups, the proportion of female participants in the control group (63.5%) was found to be significantly higher than in the PD (36.7%) and HD (33.3%) groups (p=0.002 and p=0.001, respectively). The median age (43.2 years) in the control group was significantly lower than in the PD (60.7 years) and HD (58.8 years) groups (p<0.001 and p<0.001, respectively). There was no statistically significant difference between the ratios of female participants and the PD and HD groups’ median age values (p=0.704). The median body mass index of the patients in the PD group was significantly higher than that of the HD group (p=0.039). The rates of diabetes mellitus in the HD group (48.89 vs. 26.53%, p=0.025) and hypertension in the PD group (93.88% vs. 80%, p=0.044) were significantly higher than in the other group. No statistically significant difference was found in other comparisons between PD and HD groups (p>0.05) (Table 1). The annual number of peritonitis attacks of the patients in the PD group was a minimum of 0 and a maximum of 4 (median 0).

The median leukocytes, neutrophils, lymphocytes, ferritin, total cholesterol, low-density lipoprotein (LDL) cholesterol, high-density lipoprotein (HDL) cholesterol, and calcium measurements in the PD group were higher than in the HD group, and the median uric acid level was lower (Table 2).

### 3.1 Vaccination preference

The vaccine type and dose distribution applied in the groups were shown in Table 3. It was determined that 51.74% of the participants were given CoronaVac, 31.34% combined, and 16.91% BioNTech vaccine. There was a statistically significant difference between the vaccine distributions of the groups (p<0.001). CoronaVac (46.72%) and combined vaccine (41.12%) were mostly preferred in the control group. CoronaVac vaccine (73.47%) was selected more than the combined (16.33%) and BioNTech (10.20%) vaccines in the PD group. CoronaVac (40%), BioNTech (35.56%) and combined (24.44%) vaccines were preferred in the HD group, respectively.

32.34% of the participants received two doses of the CoronaVac vaccine, and 31.34% received the combined vaccine. Participants in the control group mostly preferred the combined vaccine (41.12%), the participants in the PD group preferred three doses of CoronaVac vaccine (42.86%), and the participants in the HD group preferred two doses of BioNTech vaccine (26.67%). In the subgroup analysis, the rate of those who received three doses of the CoronaVac vaccine in the PD group was found to be higher than the patients in the control and HD groups. The rate of those who received two doses of the BioNTech vaccine was higher in the HD group than in the control group. The proportion of those who received the combined vaccine was higher in the control group than in the PD group (p<0.05).

Regardless of the vaccine type, there was a significant difference between the distributions of the total vaccine dose numbers of the groups (p=0.008) (Table 3). 48.76% of the participants received three doses, 45.77% received two doses, and 5.47% received a single dose. Two doses (48.59%) in the control group, three doses (59.18%) in the PD group, and two doses (46.67%) in the HD group were preferred more frequently. In the subgroup analysis, the rate of those who received a single-dose vaccine was higher in the HD group than in the control group (p<0.05).

### 3.2 Antibody responses

The median time between the last dose of vaccine and antibody measurement in all participants was 2.46 months (min:max 0.16-8.38). This time was determined as 3.22 (min:max 0.16-8.38) in the control group, 2.37 (min:max 0.79-6.01) in the PD group, and 1.84 (min:max 0.46-6.11) in the HD group. A statistically significant difference existed between the groups’ antibody responses to the vaccine (p=0.009). The rate of those with insufficient antibody response in the control group was lower than those in the HD group, and the rate of those with a negative response was lower than in the PD group (p<0.05). Antibody responses to the vaccine were significantly different according to the participants’ vaccine type and dose preferences (p<0.001). Compared to those in the CoronaVac or BioNTech groups, those receiving the combined vaccine had a higher rate of complete responders and a lower rate of insufficient responders (p<0.05). Compared to those who preferred one or two doses, the complete response rate was higher, and the insufficient response rate was lower in those who chose three vaccine doses. The rate of those with suboptimal response in those who preferred two vaccine doses was higher than those who decided on three doses. The rate of negative responders who chose a single-dose vaccine was higher than those who preferred two or three doses (p<0.05) (Table 4).

Antibody responses to the vaccine were statistically significantly different according to the dialysis patients’ vaccine type (p=0.006) and dose (p=0.005) preferences. Compared with patients in the CoronaVac group, patients in the combined group had a higher complete response rate and a lower suboptimal response rate. The insufficient response rate was lower in patients who preferred three doses than in patients who chose one or two. The negative response rate was also higher in those who preferred a single-dose vaccine than those who preferred two doses (p<0.05) (Table 5).

### 3.3 Post-vaccine COVID-19 infection

One person in the control group received two vaccine doses at an interval of one month. He became infected with COVID-19 23 days after the second dose of the vaccine, and three months later, he received the third dose of the vaccine. The antibody level was >250 U/mL. Twenty-five subjects in the control group, six patients in the PD group, and four in the HD group had COVID-19 infection median 6.07 (min-max: 1.74-11.76), 6.24 (min-max: 2.46-10.22) ve 7.22 (min-max: 5.19-10.41) months after the last vaccine dose, respectively.

The post-vaccine COVID-19 infection rate was 24.29% (n: 26) in the control group, 12.24% (n: 6) in the PD group, and 8.88% (n: 4) in the HD group (p=0.038, Pearson Chi-square test). Only the control group had a significantly higher incidence of post-vaccine COVID-19 infection than the HD group (p=0.029). In three groups, no significant difference was observed between the antibody responses of people with and without COVID-19 (Table 6). Twenty-six people in the control group were treated without hospitalisation. Of six patients with COVID-19 infection in the PD group, 33.33% were hospitalised, and 16.67% were treated in the intensive care unit due to the need for respiratory support and died. On the other hand, 50% of four patients in the HD group were hospitalised, and 25% were treated in the intensive care unit due to the need for respiratory support and died.

### 3.4 Analysis of risk factors for post-vaccination COVID-19 infection in dialysis patients

The determining risk factors for the development of COVID-19 infection were studied with multivariate retrospective stepwise elimination logistic regression analyses. All variables with p<0.25 in univariate analyses were included in the regression models as candidate risk factors. Only variables with a multicollinearity problem were included in the model (diabetes, hepatitis B, body mass index, hemoglobin, transferrin saturation, glucose, total cholesterol, LDL cholesterol, albumin, phosphate, CaxP product and parathyroid hormone variables). The final model results for COVID-19 infection as a result of the retrospective elimination procedure were shown in Table 7. As a result of multivariate logistic regression analysis determined that the model obtained in the final step was significant (p=0.005) and compatible with the data set (p=0.058). Diabetic dialysis patients were 7.70 times more likely to get COVID-19 infection after vaccination than non-diabetics. LDL cholesterol levels in dialysis patients did not affect the risk of contracting COVID-19 after vaccination (p=0.055). It was determined that a one-unit increase in albumin value reduced the risk of contracting COVID-19 after vaccination by 20%.

## 4 DISCUSSION AND CONCLUSIONS

This retrospective study compared vaccines’ efficacy and antibody levels in protecting against SARS-CoV-2 after inactivated (CoronaVac) and/or nucleic acid RNA (BioNTech) vaccination in dialysis patients and healthy subjects. In our country, the inactivated vaccine (CoronaVac) was started on January 14, 2021, and the RNA vaccine (BioNTech) was started on April 2, 2021. Vaccination was first opened to high-risk groups and then gradually to all age groups. Individuals were also allowed to choose the BioNTech or CoronaVac vaccine. In our study, most of the participants had the CoronaVac (51.7%) vaccine, followed by the combined (31.3%) and BioNTech (16.9%) vaccines. The rate of CoronaVac (46.7%) and combined vaccine (41.1%) in the control group, CoronaVac vaccine (73.4%) in the PD group, and CoronaVac (40%) and BioNTech (35.5%) vaccination rates in the HD group were higher. In all cohorts, two doses of the CoronaVac vaccine (32.3%) and three doses (CoronaVac 2 doses plus BioNTech 1 dose) combined vaccine (31.3%) were preferred more. A similar trend was observed in the control and HD groups (the combined vaccine 41.1% and CoronaVac two doses 38.3%), but the PD group received at most three doses of the CoronaVac vaccine (42.8%). 48.7% of the participants had three doses, 45.7% had two, and 5.47% had a single dose. Two doses were preferred more in the control (48.5%) and HD (46.6%) groups, and three doses (59.1%) in the PD group.

After the vaccines were made available to the public during the pandemic, the primary determinant in vaccine selection was access to the vaccine, the policies and guidance of WHO, local health authorities and scientists. As time progressed and the options multiplied, people started to decide for themselves. Developing inactivated vaccines with conventional methods and having a low side-effect profile were the reasons for preference. Because the technology of RNA vaccines, set in a relatively short time, was new, at least in the field of vaccines, some people were apprehensive about the vaccine’s side effects. Parallel to the increase in the number of vaccinated individuals and the variety of vaccines, scientists began to question the following issues: how much each vaccine protects, how long is the protection period of antibody levels against the vaccine, how many doses of vaccine should be given, how long the dosing interval should be, and how the vaccine policy should be followed in previously infected individuals. Undoubtedly, the constant mutation of the virus and the inability to prevent the global spread of the epidemic was another obstacle to the effectiveness of vaccines. The above reasons were influential in the vaccine preferences of our participants. Interestingly, PD patients preferred the CoronaVac vaccine more (73.4%) in contrast to the tendency of the control and HD groups to prefer CoronaVac or combined vaccines. The main reason for their preference for inactivated vaccines may be that PD patients can isolate themselves more easily than patients treated for HD at the centre. The introduction of the BioNTech vaccine approximately 3.5 months after the CoronaVac vaccine in our country may have undoubtedly been effective in patient preferences. Participants mostly had two or three doses of the vaccine. The rate of three-dose vaccination was higher in PD patients. Those who had a single dose may not have taken the other doses because of a side effect in the first dose.

In HD patients, the uremic state is associated with premature immunological ageing, a persistent inflammatory condition, and functional defects in immune cell populations [7, 8]. Uremia also impairs and inhibits the production of neutralising antibodies (NAbs) [9]. A systematic analysis supports that HD patients have lower seroconversion and seroprotection rates after vaccination against viral respiratory diseases (H1N1, H3N2 and COVID-19) than controls [10]. In a meta-analysis, the overall immunogenicity of dialysis patients after the SARS-CoV-2 vaccination was 87% [11]. High seroconversion rates were obtained without severe adverse effects after a single dose of the BioNTech (BNT162b2) vaccine in HD patients and two doses of the mRNA-1273 Moderna vaccine in PD patients [12, 13]. SARS-CoV-2 double vaccination in HD patients is associated with a significant reduction in oxygen supplementation and overall mortality [14]. However, a systematic review of eighteen studies determined that seroconversion rates (88.5% vs 27.2%) and anti-spike antibody titers were lower in dialysis and kidney transplant patients after complete vaccination with two doses of the COVID-19 mRNA vaccine compared to the healthy population [15]. Overall, studies suggest that PD and HD patients and transplant recipients after two doses of the BioNTech or vector-based (ChAdOx1 nCoV-19, Oxford-AstraZeneca) vaccines have an inferior antibody response and a lower robust response rate to COVID-19 vaccines compared to healthy controls, with a faster decline in antibody titers over time [16-18]. Population differences, vaccine and dosage settings, immune response definitions, and antibody detection time points may influence the occurrence of lower and delayed antibody responses following COVID-19 vaccination in studies [19, 20]. In our study, the complete response rates (>250 U/ml) according to the antibody level were 71.0% in the control group, 61.2% in the PD group, and 57.7% in the HD group. The number of patients with insufficient response and no response to the vaccine was 5 and 3 in the HD group and three each in the PD group, respectively. Of these patients, 2 had three doses, 7 had two doses, 1 had a single dose of CoronaVac, 2 had a single dose and 1 had a double dose of BioNTech.

The seroconversion rates and antibody levels are lower with two doses of inactivated vaccine in dialysis patients than in healthy controls [21]. After two doses of inactivated whole-virus SARS-CoV-2 vaccine (CoronaVac), the receptor binding protein (anti-RBD) IgG antibody levels was significantly higher in healthy controls than in dialysis patients but was comparable between HD and PD groups [22]. In many studies, CoronoVac or mRNA vaccinations induced adaptive immune response in PD and HD patients, with no difference in antibody levels between modalities [6, 23-25]. Other studies show that the mRNA vaccine has a higher seropositivity rate and quantitative antibody levels than an inactivated vaccine in dialysis patients, but both vaccines protect against symptomatic infection when seropositivity is achieved [26, 27]. In a large HD cohort, two doses of the BNT162b2 (Pfizer-BioNTech) vaccine were more effective in preventing SARS-CoV-2 infection (42.6% vs 15.0%) and COVID-19-related deaths (90.4% vs 64.8%) compared to CoronaVac (Sinovac) vaccine [28].^8^ PD patients respond more strongly to two doses of the BNT162b2 vaccine than HD and kidney transplant patients [29]. In our study, antibody levels were measured in all participants median of 2.46 months after the last dose of vaccine. Complete plus suboptimal antibody response rates were higher in healthy individuals than in PD and HD patients (99.0% vs 87.7% and 84.3%, respectively). Since our study was retrospective, we could only look at a single antibody level measured median 2.46 months after the last vaccine dose. As expected, the antibody response was better in healthy individuals. There was no significant difference between PD and HD patients.

The combined vaccine or three doses provided a higher complete antibody response than the other options in our cohort. Regardless of the vaccine type, an increase in doses was positively associated with the complete response rate. While the complete antibody response was 83.6% in those who received three doses, it was lower in those with a single (45.4%) or two (48.9%) doses. A study including 13,759 maintenance dialysis patients (17% unvaccinated and 83% vaccinated) revealed that although one dose of the COVID-19 mRNA vaccine provided weak protection, two doses effectively prevented SARS-CoV-2 infection and associated severe outcomes [30]. The blunted cellular response to vaccines, the incomplete and delayed humoral response and the decrease in antibody titers over time in dialysis patients support the need for additional doses or the use of different vaccines or combinations of vaccines [31-33]. In HD and PD patients, antibody levels increased significantly after the third dose of the BNT162b2 vaccine [34]. The third dose of the BNT162b2 vaccine enables 40% of patients to switch from null or partial protection after two doses to complete protection. Many studies show increased humoral response with a third booster dose to HD patients, but the exact timing and dosing schedule still need to be determined [35, 36]. A non-negligible proportion of dialysis patients still have low titers after a third dose and may benefit from a fourth booster dose [37]. During our study, individuals were given a maximum of three vaccine doses. Due to the continuation of waves due to various variants in our country during the pandemic, fourth and fifth doses of inactivated or mRNA BNT162b2 vaccine were administered to individuals, including dialysis patients.

The relatively low seroconversion rates and gradually decreasing antibody titers in dialysis patients have prompted the search for other strategies to improve the efficacy of vaccines. Several studies have shown that different combinations of vaccines provide more effective protection against COVID-19. Administration of the third dose of the ChAdOx1 nCoV-19 vaccine (AZD1222, Oxford University-AstraZeneca) increased immunogenicity in PD and HD patients whose immune response was inadequate with two doses of inactivated vaccine (CoronaVac) [38]. In our study, the highest response rate was obtained with CoronaVac plus BioNTech (96.8%), followed by BioNTech (70.8%) and CoronaVac (45.1%) vaccines. The response was suboptimal in only two patients in the CoronaVac plus BioNTech group. Suboptimal antibody response rates in those vaccinated with CoronaVac or BioNTech were 44.2% and 20.5%, respectively. Factors associated with a non-response to the vaccine or lower antibody levels in dialysis patients included older age, diabetes mellitus, immunosuppressive therapy, lower lymphocyte count, lower serum albumin and high C-reactive protein levels in different studies [15, 32, 39-42].

Apart from vaccine type, number of vaccine doses and dose intervals, a history of pre- or post-vaccine COVID-19 infection may also affect antibody response. HD patients develop a detectable humoral response for at least seven months after infection with COVID-19 [43]. HD patients recovering from severe COVID-19 may have a sustained anti-spike protein seroconversion even one year after diagnosis [44]. Many studies show that dialysis patients with COVID-19 have a higher antibody response rate and/or higher anti-spike titers after mRNA vaccines than those without COVID-19 [44-49]. The neutralising capacity of SARS-CoV-2 antibodies is significantly increased in patients with SARS-CoV-2 infection, even after a single vaccination dose [50]. In our cohort, one healthy person became infected between the 2nd and 3rd doses of vaccines. Another 35 subjects (23.3% in the control group, 12.2% in the PD group and 8.8% in the HD group) became infected with COVID-19 at a median of 6.27 months (range 1.74-11.76) after the last dose of vaccine. The post-vaccine infection rate was higher in the control group than in the PD and HD groups. The antibody response rates of participants with and without COVID-19 were comparable. In the subgroup analysis, six infected patients in the PD group preferred two or three doses of the CoronoVac vaccine. This rate was significantly higher than the 26 healthy individuals infected (CoronaVac 46.2%, combined 42.3% and BioNTech 11.5%, p=0.042). While the complete response rate was 76.9% and the suboptimal response rate was 23.1% in healthy individuals, the complete, suboptimal, and insufficient response rates were 33.3% each in PD patients (p=0.018). However, while healthy individuals recovered without severe problems, the need for hospitalisation and mechanical ventilation-intensive care and mortality (total 2, one PD and HD patient) were observed in some dialysis patients. Multivariate regression analysis revealed that the presence of diabetes in dialysis patients increased the probability of contracting COVID-19 infection 7.70 times after vaccination compared to those without diabetes. In addition, a one-unit increase in albumin value reduced the risk of contracting COVID-19 after vaccination by 20%. In a study comparing NAbs activity (surrogate virus neutralisation test, iFlash-2019-nCoV Nab) and TAb titer (total anti-SARS-CoV-2: neutralising and non-neutralizing anti-S antibodies) in the sera of dialysis patients, TAb titer rates were higher than NAbs titers four weeks after the second dose of vaccine in patients previously infected with SARS-CoV-2 and after the third dose in patients without a history of COVID-19. NAbs values increased after the third dose in non-COVID-19 patients, but lower production in infected patients was seen four weeks after the second dose compared to the first dose of the vaccine [9]. Our expectation of better antibody response in those with previous SARS-CoV-2 infection may not be valid for all patients. In the studies discussed above, many factors such as vaccine type, vaccine dose, positive immune response criteria, antibody detection times, race and ethnicity may have affected the immune response. Despite the recommendation of three doses for COVID-19 naive HD patients and two doses for previously infected patients, we should be aware that two doses may provide adequate protection in some patients. In contrast, even three doses may result in poor protection in others. This study did not have detailed data on the vaccine preferences of the groups and the side effects of the vaccines. Since the study was retrospective, the exact level of antibody titers above 250 U/mL was unknown.

According to the hospital COVID-19 case records, the peaks of COVID-19 waves in our city between March 2020 and March 2022 were as follows: the peak of the first wave in April 2020, the peak of the second wave in November 2020 (original alpha variant, October-December 2021), the peak of the third wave (delta variant) in April 2021 (March-May 2021), the peak of the fourth wave (delta variant) in October 2021 (September-December 2021) and the peak of the fifth wave (omicron variant) in February 2022 (December-March 2022). Cellular and humoral immune responses to SARS-CoV-2 in the dialysis population are variable. Response rates to the COVID-19 vaccine in dialysis patients range from 29.6% to 96.4% [39]. To whom and when booster doses should be given is still being determined. Similar to ours, vaccination preferences in dialysis patients showed marked heterogeneity in many studies. Due to the high risk of contracting SARS-CoV-2 and developing severe COVID-19 in dialysis patients, SARS-CoV-2 vaccination schedules must be personalised. In addition to personal protection, vaccination will always be a priority, not only in COVID-19 but also in other potential viral infection outbreaks that may occur in the future. With the International Health Regulations (IHR) Emergency Committee report on the COVID-19 pandemic, held on 4 May 2023, WHO recommended a transition to long-term management despite uncertainties in the potential evolution of SARS-CoV-2. WHO has declared that although COVID-19 no longer constitutes a public health emergency of global importance, it will continue to be a health problem. The dialysis population will be particularly vulnerable to infection with the new variants because antibody levels decrease over time in patients who respond after vaccination. Our pandemic experience justifies that sporadic cases will continue in dialysis patients, so annual COVID-19 boosters and flu shots must be repeated in risky months.

## Data Availability

All data produced in the present study are available upon reasonable request to the authors

